# Analysis of Social Determinants of Hypertension Outcomes among Asian Americans in Los Angeles

**DOI:** 10.1101/2023.10.17.23296571

**Authors:** Megan R. Yu, Haoyuan Liu, Madyllen Kung, Caitlyn Tran, Shameek Mitra, Preston Dang, Benjamin K. Woo

**Author notes:** Denotes equal contributions (Megan R. Yu, Haoyuan Liu, Madyllen Kung, Caitlyn Tran, Shameek Mitra).

## Abstract

Asian Americans are a diverse and significant group within the United States and encompass a wide range of social demographics. Research on social determinants of hypertension within this population is limited, despite a notable burden of illness. Asian American attendees at health fairs held in Monterey Park and Rosemead, California in Los Angeles County were surveyed on various social demographics and screened for presence of hypertension outcomes. Logistic regression modeling was employed to determine major demographic contributors to the prevalence of prehypertensive or higher outcomes, ultimately finding that health fair location was a significant predictor (OR: 3.41; 95% CI: 1.54, 7.58; P-Value: 0.003). Further analysis of only attendees exhibiting prehypertensive or higher outcomes showed a significant distribution in the conditional distribution of education levels between Monterey Park and Rosemead attendees. Study findings uncover further areas for research into both Asian American and Chinese American populations and contribute to an overall gap in research within these demographic groups.

## Introduction

### Asian American Presence and Wide Variation in Social Demographics

Based on reporting from the Census Bureau as of May 2023, around 24 million Asian Americans reside in the United States [1]. The Asian American population is very diverse, representing over 20 countries across South, East, and Southeast Asia. Chinese Americans are the most represented origin group, accounting for about one-quarter of the overall Asian American population. About half of all Asian Americans live in the Western states, with California being home to the most Asian Americans of any other US state (6.7 million) as of 2019 [2]. Los Angeles County in California is home to both the largest population as well as the fastest growing population of Asian Americans in the country between 2000 and 2010 based on Census data [3]. Finally, notable diversity has been previously observed in the settlement patterns of Asian Americans across numerous communities within Los Angeles County, where enclaves of different Asian subgroups differ based on geographic location within the county [4].

The model minority myth is a misleading social assumption that professes a monolithic narrative of high income and educational attainment for Asian Americans, thus minimizing the varied social experiences of different Asian American subgroups [5]. While the Asian American population generally does well on measures of economic well-being compared to the entire U.S. population, such as median annual household income, there is wide variation among individual Asian subgroups. As of 2019, the median household income for all Asian subgroups was $86,000. However, Indian Americans were found to have the highest annual income of all Asian American subgroups ($119,000), while Burmese Americans were found to have the lowest ($44,000). A similar trend is observed within measures of educational attainment, according to a report by the Pew Research Center in 2019. While 54% percent of Asian Americans 25 and older hold a bachelor’s degree or higher, the highest percentage was seen among Indian Americans at 75% and the lowest observed among Bhutanese Americans at 15% [6].

### Limited Research on Social Determinants of Hypertension among Asian Americans

Previous research has shown significant associations between favorable outcomes of hypertension and higher incomes [7], presence of health insurance [8], and frequency of doctor’s visits [9]. Many studies have also shown consistent evidence of a relationship between daily physical activity and lowered prevalence of hypertension [10]. Although previous reporting from the National Center for Health Statistics showed a significant association between hypertension prevalence and lower education attainment among Asian adults in the United States from 2011-2012 [11], research on social determinants of hypertension among Asian Americans is limited overall. Given that heart disease was the second-leading cause of death amongst both Asian men [12] and women [13] in the United States in 2018 according to the Centers for Disease Control and Prevention and hypertension has been shown to disproportionally affect different Asian subgroups [14], it is crucial to further elucidate the effects of social determinants of hypertension among Asian Americans as a whole and within Los Angeles county.

Through this study, we seek to model various social determinants of hypertension among Asian Americans: education, income, insurance, regular doctor’s visits, and daily physical activity. Given the geographic diversity of Asian American demographics in Los Angeles County, we also explored location as a potential predictor of hypertension within this population. Results were obtained through logistic regression and Chi-Square analysis using data collected from free community health fairs held within the greater Los Angeles area organized by a student-run health service organization, Asian Pacific Health Corps at UCLA (University of California, Los Angeles), between November 2019 and January 2020.

## Methods

Convenience samples were performed at free health fairs held in Monterey Park (MPHF) and Rosemead (FDHF), California for this retrospective, secondary analysis. A total of 318 AAPIs were included in the working sample, including 150 AAPIs from MPHF and 168 from FDHF. All other demographic predictors of interest, including ethnicity, education, income, health insurance status, presence of regular doctor’s visits, and daily physical activity, were recorded through a voluntary paper survey offered at the entrance of each fair. Gender, age, systolic and diastolic blood pressure, and body mass index (BMI) values were collected at screening stations inside the fair if attendees elected to undergo these screenings. No personally identifiable information was recorded from attendees; however, an arbitrary number was assigned to each attendee upon entry to link data at all points of data collection.

As the primary outcome of interest, blood pressure values were classified using ranges provided by the American Heart Association (AHA) [15]. For analysis, values for blood pressure were aggregated into a binary outcome based on the category ranges provided by the AHA. All values in the “Normal” range based on AHA guidelines and lower were classified as “Normal or Lower”; conversely, all values considered by the AHA to be in the “Prehypertension” range and higher were classified as “Prehypertensive or Higher.” Ethical considerations for this project were satisfied through the University of California, Los Angeles Institutional Review Board (Protocol ID: PRE#20-003027IRB) through designation as non-human subject research.

All statistical analyses and tabulations were performed using Stata 17.0 (StataCorp. 2021. Stata Statistical Software: Release 17. College Station, TX: StataCorp LLC.). Statistical methods included categorical inferential procedures such as Chi-Square and Fisher’s Exact testing as well as logistic regression modeling. Forward selection was used to test predictors based on p-values from individual simple logistic regression models between each predictor and presence of hypertension outcome for inclusion in the final adjusted model. Appropriate conditions and diagnostic procedures were satisfied for each statistical procedure.

## Results

Table 1 summarizes the demographic characteristics of the overall Asian American health fair attendee sample and within each health fair. An overwhelming proportion of attendees reported being of Chinese ethnicity, though a large variety of other Asian ethnicities were represented in much smaller proportions. Most Asian American attendees were female with an overall median age of 66 and interquartile range of 13 years, though the majority of data were missing for these variables. About half of overall Asian American attendees reported college or high school as their highest level of education and having an annual income of less than $20,000. About two-thirds of all Asian American attendees reported having health insurance and visiting their doctor regularly and about 40% of all Asian American attendees reported exercising between 30 and 60 minutes per day. No significant differences in the conditional distributions of education, income, health insurance status, regular doctor’s visits, and daily physical activity were observed between MPHF and FDHF based on Chi-Square testing.

**Table 1:**
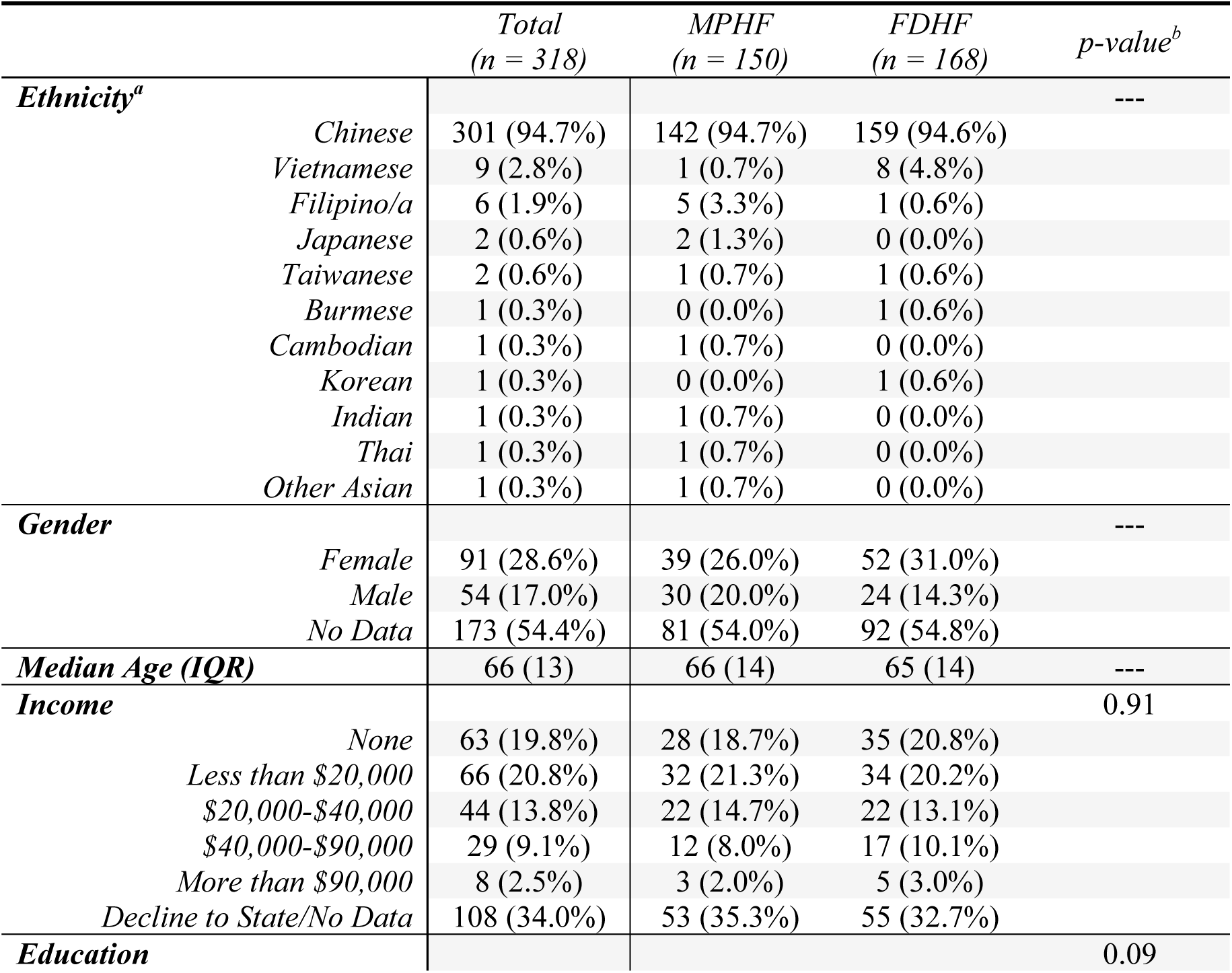
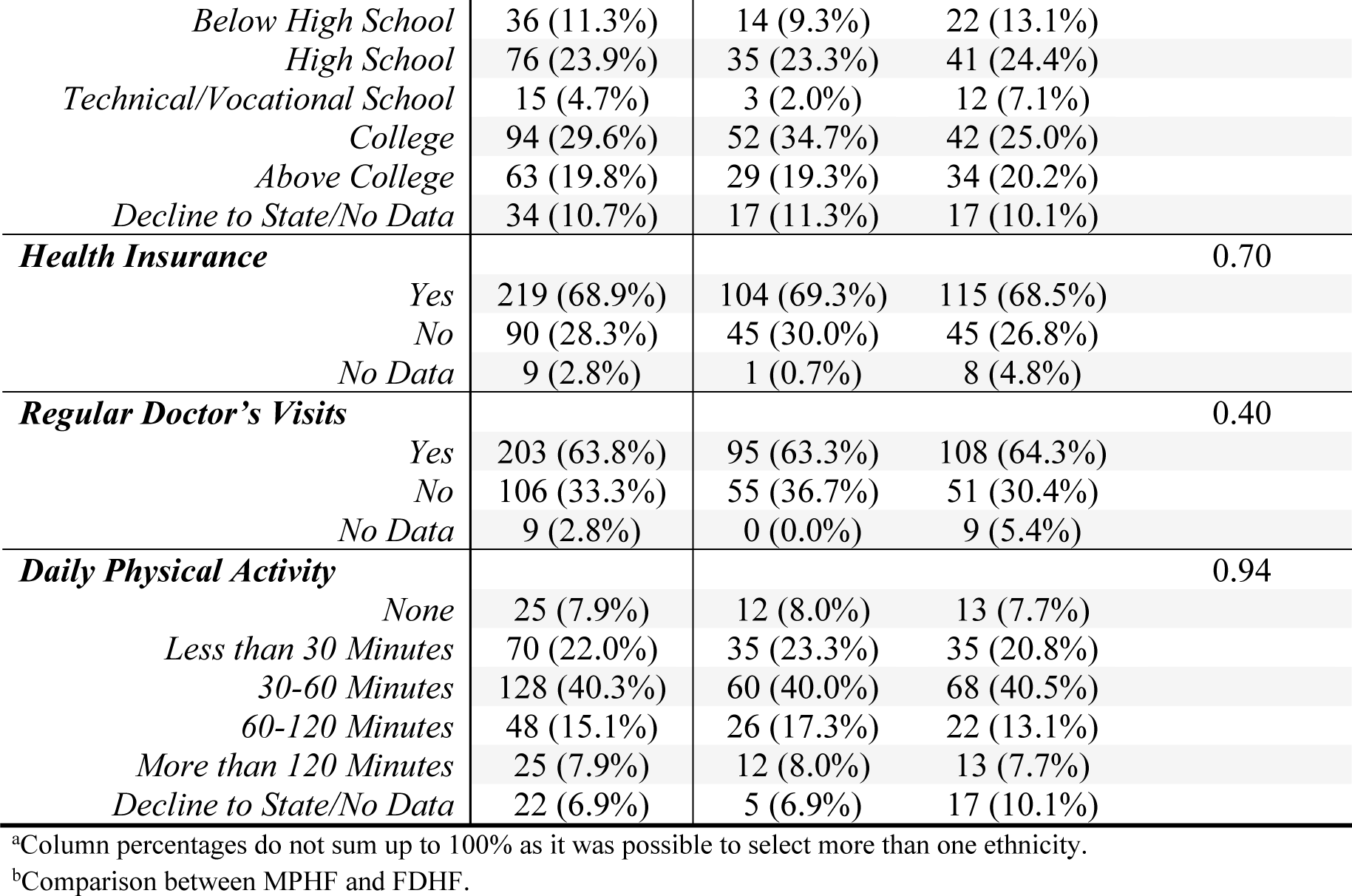
Demographics of Asian American Health Fair Attendees.

Table 2 displays results from simple logistic regression (SLR) modeling of prehypertensive outcomes and above with each individual demographic predictor. Health fair location was found to be a significant predictor of attendee odds of experiencing prehypertensive outcomes and above. No other demographic factors were found to be significant and were thus excluded from the final adjusted model.

**Table 2:**
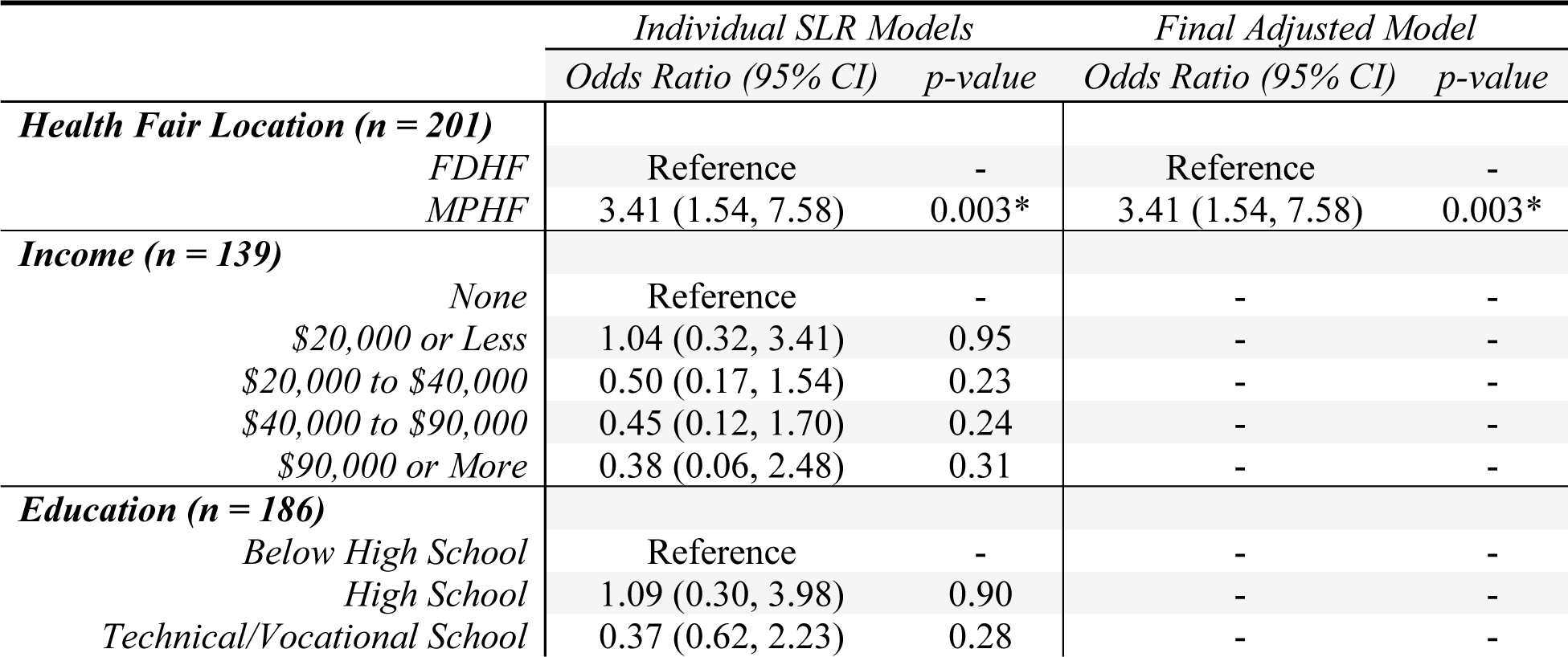
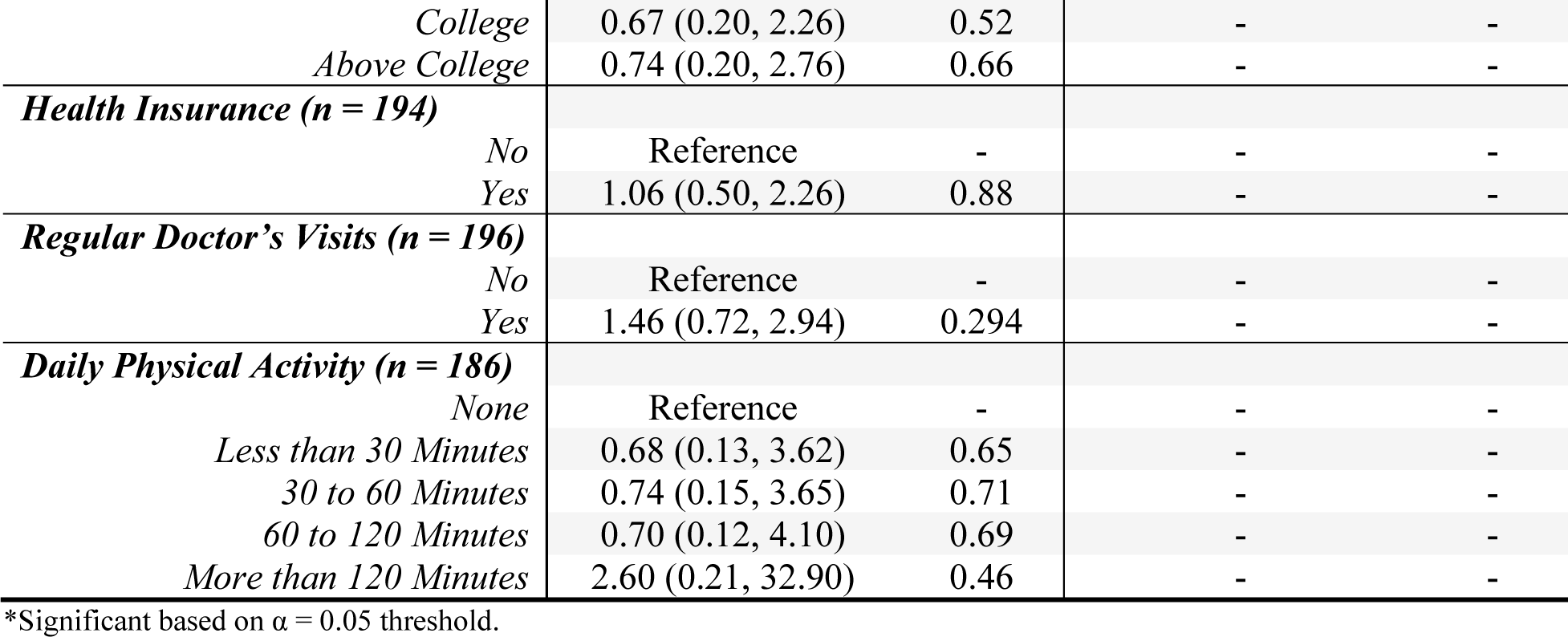
Logistic Regression Output for Prehypertensive Outcomes and Above Asian American Attendees.

|  | *Individual SLR Models* |  | *Final Adjusted Model* |  |
| --- | --- | --- | --- | --- |
|  | *Odds Ratio (95% CI)* | *p-value* | *Odds Ratio (95% CI)* | *p-value* |
| **Health Fair Location (n = 201)** |  |  |  |  |
| *FDHF* | Reference | - | Reference | - |
| *MPHF* | 3.41 (1.54, 7.58) | 0.003* | 3.41 (1.54, 7.58) | 0.003* |
| **Income (n = 139)** |  |  |  |  |
| *None* | Reference | - | - | - |
| *\$20,000 or Less* | 1.04 (0.32, 3.41) | 0.95 | - | - |
| *\$20,000 to \$40,000* | 0.50 (0.17, 1.54) | 0.23 | - | - |
| *\$40,000 to \$90,000* | 0.45 (0.12, 1.70) | 0.24 | - | - |
| *\$90,000 or More* | 0.38 (0.06, 2.48) | 0.31 | - | - |
| **Education (n = 186)** |  |  |  |  |
| *Below High School* | Reference | - | - | - |
| *High School* | 1.09 (0.30, 3.98) | 0.90 | - | - |
| *Technical/Vocational School* | 0.37 (0.62, 2.23) | 0.28 | - | - |
| *College* | 0.67 (0.20, 2.26) | 0.52 | - | - |
| *Above College* | 0.74 (0.20, 2.76) | 0.66 | - | - |
| **Health Insurance (n = 194)** |  |  |  |  |
| *No* | Reference | - | - | - |
| *Yes* | 1.06 (0.50, 2.26) | 0.88 | - | - |
| **Regular Doctor's Visits (n = 196)** |  |  |  |  |
| *No* | Reference | - | - | - |
| *Yes* | 1.46 (0.72, 2.94) | 0.294 | - | - |
| **Daily Physical Activity (n = 186)** |  |  |  |  |
| *None* | Reference | - | - | - |
| *Less than 30 Minutes* | 0.68 (0.13, 3.62) | 0.65 | - | - |
| *30 to 60 Minutes* | 0.74 (0.15, 3.65) | 0.71 | - | - |
| *60 to 120 Minutes* | 0.70 (0.12, 4.10) | 0.69 | - | - |
| *More than 120 Minutes* | 2.60 (0.21, 32.90) | 0.46 | - | - |

\*Significant based on $\alpha = 0.05$ threshold.

Table 3 displays conditional distribution counts of all demographic predictors of interest and results of Chi-Square testing among only MPHF and FDHF attendees experiencing prehypertensive or higher outcomes. A significant difference was observed between MPHF and FDHF attendees only with respect to education and no other predictors.

**Table 3:**
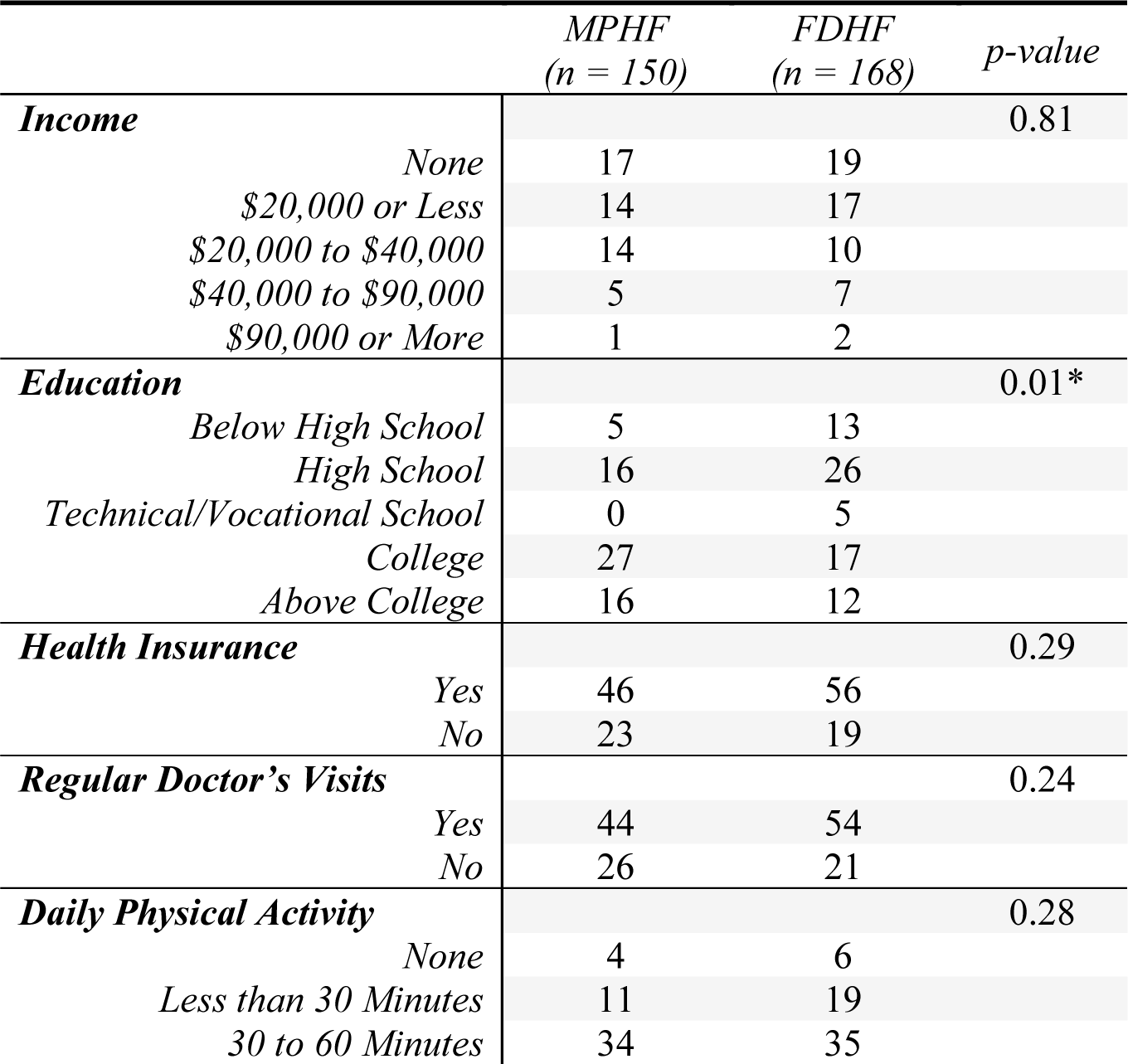
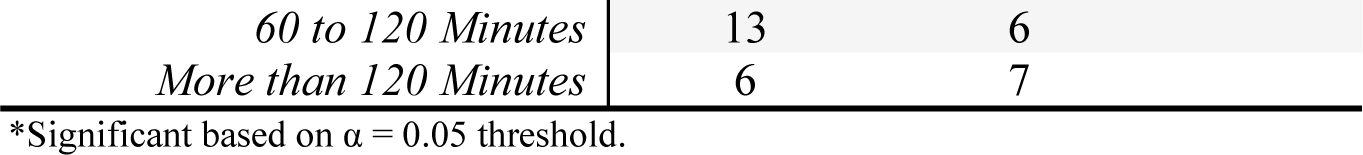
Conditional Count Distributions of Demographic Predictors of Interest among Asian American Attendees with Prehypertensive or Higher Outcomes at MPHF and FDHF.

|  | *MPHF*<br>(n = 150) | *FDHF*<br>(n = 168) | *p-value* |
| --- | --- | --- | --- |
| **Income** |  |  | 0.81 |
| *None* | 17 | 19 |  |
| *\$20,000 or Less* | 14 | 17 | |
| *\$20,000 to \$40,000* | 14 | 10 | |
| *\$40,000 to \$90,000* | 5 | 7 | |
| *\$90,000 or More* | 1 | 2 | |
| **Education** |  |  | 0.01* |
| *Below High School* | 5 | 13 |  |
| *High School* | 16 | 26 |  |
| *Technical/Vocational School* | 0 | 5 |  |
| *College* | 27 | 17 |  |
| *Above College* | 16 | 12 |  |
| **Health Insurance** |  |  | 0.29 |
| *Yes* | 46 | 56 |  |
| *No* | 23 | 19 |  |
| **Regular Doctor's Visits** |  |  | 0.24 |
| *Yes* | 44 | 54 |  |
| *No* | 26 | 21 |  |
| **Daily Physical Activity** |  |  | 0.28 |
| *None* | 4 | 6 |  |
| *Less than 30 Minutes* | 11 | 19 |  |
| *30 to 60 Minutes* | 34 | 35 |  |

|  |  |  |
| --- | --- | --- |
| 60 to 120 Minutes | 13 | 6 |
| More than 120 Minutes | 6 | 7 |

\*Significant based on $\alpha = 0.05$ threshold.

## Discussion

### Overwhelming Representation of Chinese Americans

Our analytical sample featured an overwhelming representation of Chinese American individuals, accounting for over 90% of overall Asian American attendees and within attendees at each health fair. Los Angeles county is home to the highest number of Chinese Americans of any other county in the United States. Between 2000-2010, the growth rate of the Chinese American population (20%) outpaced that of the county overall (3%) [3]. In particular, the communities of Monterey Park and Rosemead within Los Angeles County are home to the highest and sixth-highest proportions of ethnic Chinese individuals based on data from the 2000 Census [16]. While this observation introduces heavy restrictions on the generalizability of our conclusions, increased representation of Asian Americans in research is introduced and interesting new avenues of further research are presented. Given the geographic diversity of the Asian American population within Los Angeles County [4], further research into these communities would help provide a more complete picture of the Asian American experience overall. In contrast, research could also be focused more into other areas of higher Chinese American presence within greater Los Angeles [16]. Either focus area would help fill crucial needs for Asian American representation within research.

### Influence of Location and Education Level on Prehypertensive and Above Outcomes

Despite no significant differences in the conditional distribution of all demographic covariates of interest, MPHF Asian American attendees exhibited over three times higher odds than FDHF Asian American attendees of experiencing a prehypertensive or above outcome based on multivariable logistic modeling. Further analysis among only Asian American attendees that recorded a prehypertensive or higher then showed a significant difference in the conditional distribution of educational level between MPHF and FDHF Asian American attendees. Based on reporting by the Pew Research Center in 2021, over half of Asian Americans in the United States possess at least a college degree [2], placing the Monterey Park community at an above average proportion of college-educated Asian Americans based on the results of this survey. While post-secondary degrees hold higher long-term value and higher levels of employment overall [17], occupational stress and high job strain has been shown to be associated with increased ambulatory blood pressure and sustained hypertension [18]. Within the context of the Asian American population of Monterey Park, occupational stress may be a contributing factor to the high prevalence of hypertension. Further analysis can consider specific occupations or working experiences of Asian Americans as potential links to the higher prevalence of hypertensive outcomes.

### Limitations

Many limitations are present in this analysis. Analysis of community health fair attendees decreases the external validity of the results of this study to represent the overall Asian American population of greater Los Angeles and its many diverse ethnic subgroups, given the demographics of these populations may be more likely to account for higher prevalences of various health conditions and lower socioeconomic attainment. A cross-sectional survey design does not account for consistency in both the demographics and hypertensive outcomes of health fair attendees as these classifications may change with time. Future studies therefore will greatly benefit from a cohort design for individual attendees, provided they continue visiting fairs in the future, or an ecological design to encompass the general populations at these fairs over time.

Limitations also exist in the inability to validate information provided by health fair attendees. Collection of information relied solely on self-reporting, which could yield inaccuracies because of response and information bias. Finally, non-consideration of other vital characteristics of the study population, such as pre-existing health conditions (genetics, etc.) or other lifestyle practices (smoking, diet, etc.), present areas of possible confounding in the resulting trends and associations of health outcomes among attendees and should be accounted for in future research.

## Conclusion

The dynamics of the Asian American population are complex and nuanced. Lack of representation of Asian Americans in research works to cloud the visibility of the burdens of chronic health conditions such as hypertension. Given the significant presence of this population in the United States, expansion of research is crucial to better understanding proper steps of tailoring health interventions and care for these communities.

## Data Availability

All data produced in the present study are available upon reasonable request to the authors.

